# Antibody longevity and cross-neutralizing activity following SARS-CoV-2 wave 1 and B.1.1.7 infections

**DOI:** 10.1101/2021.06.07.21258351

**Authors:** Liane Dupont, Luke B. Snell, Carl Graham, Jeffrey Seow, Blair Merrick, Thomas Lechmere, Sadie R. Hallett, Themoula Charalampous, Adela Alcolea-Medina, Isabella Huettner, Thomas J. A. Maguire, Sam Acors, Nathalia Almeida, Daniel Cox, Ruth E. Dickenson, Rui Pedro Galao, Jose M. Jimenez-Guardeño, Neophytos Kouphou, Marie Jose Lista, Suzanne Pickering, Ana Maria Ortega-Prieto, Harry Wilson, Helena Winstone, Cassandra Fairhead, Jia Su, Gaia Nebbia, Rahul Batra, Stuart Neil, Manu Shankar-Hari, Jonathan D. Edgeworth, Michael H. Malim, Katie J. Doores

## Abstract

As SARS-CoV-2 variants continue to emerge globally, a major challenge for COVID-19 vaccination is the generation of a durable antibody response with cross-neutralizing activity against both current and newly emerging viral variants. Cross-neutralizing activity against major variants of concern (B.1.1.7, P.1 and B.1.351) has been observed following vaccination, albeit at a reduced potency, but whether vaccines based on the Spike glycoprotein of these viral variants will produce a superior cross-neutralizing antibody response has not been fully investigated. Here, we used sera from individuals infected in wave 1 in the UK to study the long-term cross-neutralization up to 10 months post onset of symptoms (POS), as well as sera from individuals infected with the B.1.1.7 variant to compare cross-neutralizing activity profiles. We show that neutralizing antibodies with cross-neutralizing activity can be detected from wave 1 up to 10 months POS. Although neutralization of B.1.1.7 and B.1.351 is lower, the difference in neutralization potency decreases at later timepoints suggesting continued antibody maturation and improved tolerance to Spike mutations. Interestingly, we found that B.1.1.7 infection also generates a cross-neutralizing antibody response, which, although still less potent against B.1.351, can neutralize parental wave 1 virus to a similar degree as B.1.1.7. These findings have implications for the optimization of vaccines that protect against newly emerging viral variants.

## Introduction

Neutralizing antibodies against the Spike glycoprotein of severe acute respiratory syndrome coronavirus 2 (SARS-CoV-2) are important in protection from re-infection and/or severe disease.^1-6^ Vaccines that protect against COVID-19 have been rapidly developed, and an important component of these vaccines is the elicitation of neutralizing antibodies that bind the SARS-CoV-2 Spike protein, in particular the receptor binding domain (RBD). A major challenge in controlling the COVID-19 pandemic will be elicitation of a durable neutralizing antibody response that also provides protection against SARS-CoV-2 emerging variants. Whilst the kinetics and correlates of the neutralizing antibody response have been extensively studied in the early phase following SARS-CoV-2 infection,^7-12^ information on durability and long-term cross-reactivity of the antibody response against SARS-CoV-2 following infection and/or vaccination is limited due to its recent emergence in the human population and large-scale COVID-19 vaccination only being initiated in December 2020.

We have previously studied the antibody response in SARS-CoV-2 infected healthcare workers and hospitalized individuals in the first 3 months following infection using longitudinal samples^8^. We showed that the humoral immune response was typical of that following an acute viral infection where the sera neutralizing activity peaked around 3-5 weeks post onset of symptoms (POS) and then declined as the short-lived antibody-secreting cells die.^3^ However, it remained to be seen whether the neutralizing antibody response would continue to decline after the first 3 months POS or reach a steady state. In the absence of current long-term COVID-19 vaccine follow-up, knowledge of the longevity of the neutralizing antibody response acquired through natural infection in wave 1 of the COVID-19 pandemic at late timepoints (up to 10 months POS) may provide important indicators for the durability of vaccine induced humoral immunity.

SARS-CoV-2 variants encoding mutations in Spike have been identified and include B.1.1.7 (initially reported in the UK),^13^ P.1 (first reported in Brazil) and B.1.351 (first reported in South Africa)^14^ which have been associated with more efficient transmission.^15-17^ Mutations of particular concern for vaccine immunity are those present in the receptor binding domain (RBD) of Spike which is a dominant target for the neutralizing antibody response.^18^ Despite B.1.1.7, P.1 and B.1.351 showing increased resistance to neutralization by convalescent and vaccinee sera collected at the peak of the antibody response,^19-29^ cross-neutralizing activity has been observed, albeit at a lower magnitude. In contrast, complete loss of neutralization has been observed for some monoclonal antibodies targeting specific epitopes on either the *N*-terminal domain (NTD) or RBD of Spike.^20,22,24,25,30^ Combined, these studies indicate that Spike mutations may be arising in part due to the selective pressure of neutralizing antibodies in convalescent plasma^31-33^. To counter such mutations and their attendant antigenic changes, vaccines using the Spike proteins from these variants of concern (VOCs) are under investigation.^34-37^ Whether the variant Spikes will elicit a robust neutralizing response with superior cross-neutralizing activity against parental strains and newly emerging variants has not been extensively studied.^26,38,39^ Natural infection provides an important opportunity to compare the neutralizing antibody titres and cross-neutralizing activity generated from individuals exposed to different Spike variants and will give insights into how mutations in Spike impact immunogenicity, thereby informing the design of second generation vaccine candidates based on VOCs.

In this study we set out to investigate; i) the longevity of the neutralizing and cross-neutralizing antibody response against viral variants from wave 1 infections up to 10 months POS, ii) the immunogenicity of the B.1.1.7 Spike in natural infection, and iii) cross-reactivity of sera following B.1.1.7 infection. We collected sera between 145-305 days POS from individuals infected in wave 1 that were in our original hospitalized patient and healthcare worker cohorts, as well as sera from individuals with a confirmed B.1.1.7 infection between 6-73 days POS. Following the initial decline phase, neutralization titres reached a steady state and could be detected in the majority of sera collected up to 10 months POS. We observed cross-neutralization of wild-type (Wuhan strain, WT), B.1.1.7, P.1 and B.1.351 pseudotyped viral particles for both wave 1 and B.1.1.7 sera. The B.1.351 variant showed the greatest reduction in neutralization sensitivity although the fold change in neutralization compared to WT diminished at later times POS. Importantly, B.1.1.7 infection generated neutralizing antibody titres against B.1.1.7 and WT virus that were more similar to each other than was observed for wave 1 sera, indicating maintained efficacy against previously circulating strains. Overall, these findings provide important insights into long-term immunity to SARS-CoV-2 and have implications for optimization of vaccines that protect against newly emerging viral variants.

## Results

### IgG to Spike persist for up to 10 months post onset of symptoms

Our initial study measured antibody responses in sera up to 3 months POS in hospitalized patients and healthcare workers experiencing a range of COVID-19 severity, from asymptomatic infection to requiring ECMO.^8^ Additional serum samples were collected from a subset of these individuals at time points >100 days POS when they returned to hospital as part of their routine clinical care, as well as from HCW still working at St Thomas’ Hospital. No participants had received the COVID-19 vaccine at serum collection. In total, 64 sera were collected from 38 individuals, including 16 sera collected between 145-175 days POS (TP3), 29 collected between 180-217 days (TP4), and 19 collected between 257-305 days POS (TP5). We first determined the presence of IgM and IgG against Spike, RBD and N in patient sera collected at >100 days POS (**Figure 1A-F**). OD values were measured for sera diluted at 1:50. Although the IgM response decreased to low levels against S, RBD and N at later timepoints, IgM was still detected against all three antigens in some individuals. The IgG response also decreased over time to some extent for most individuals but remained detectable at timepoints up to ∼300 days POS. Those with IgG OD values near to baseline spanned across all severity groups.

**Figure 1:**
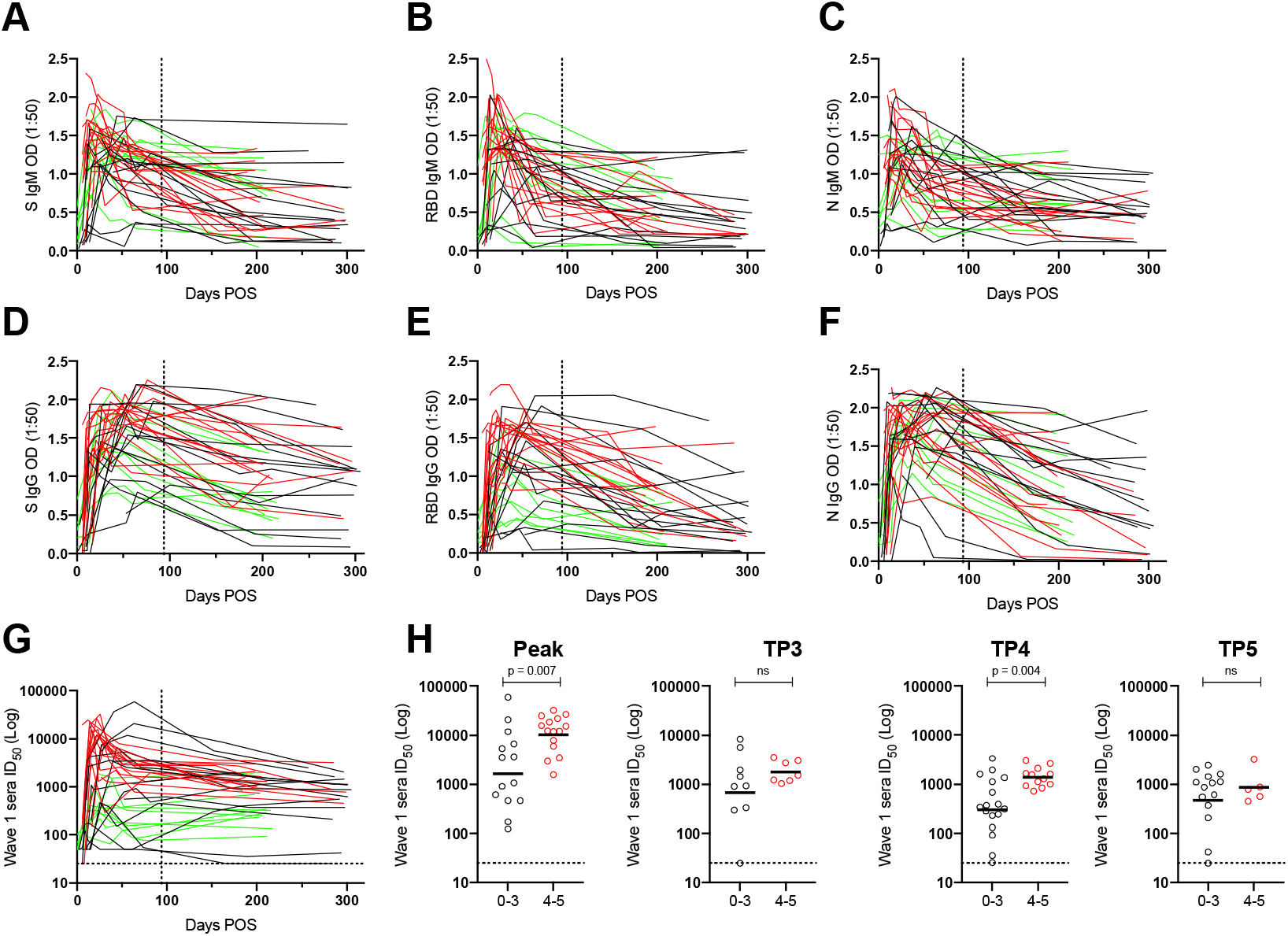
Serum Spike IgG binding and neutralizing activity is sustained up to 305 days POS. ELISA was used to assess the binding of A) IgM to Spike, B) IgM to RBD, C) IgM to N, D) IgG to S, E) IgG to RBD and F) IgG to N. Sera was diluted to 1:50 and samples were run in duplicate. The vertical dotted line indicates the time period that was studied in our original analysis of this cohort.^8^ Each line represents one individual, and they are colour coded as follows: red – severity 4-5, black – severity 0-3 and green – healthcare workers. G) Neutralization (ID_50_) measured against HIV-1 pseudotyped virus particles expressing the Wuhan Spike (wild-type, WT). The vertical dotted line indicates the latest timepoint studied in our original analysis of this cohort.^8^ H) Comparison of the mean ID_50_ between individuals experiencing 0-3 and 4-5 disease severity at different times post onset of symptoms (POS) and for the highest neutralization titre measured (Peak). Severity 0-3 is shown in black and severity 4-5 is shown in red. *p*-values were calculated using a Mann–Whitney two-sided test *U*-test. ns, not significant. The line represents the geometric mean ID_50_ for each group.

We previously used pre-COVID-19 control sera to set a threshold OD value of 4-fold above background as a cut-off for SARS-CoV-2 seropositivity.^40^ Using this cut-off, 5/45 (11.1%) and 3/19 (6.7%) of individuals had IgG below the cut-off against all three antigens (S, RBD and N) between TP3+4 and TP5, respectively. The lowest seroreactivity was observed against RBD at timepoints >145 days POS. IgG to N has been used as an indicator of previous SARS-CoV-2 infection when studying COVID-19 vaccine responses.^41,42^ However, at >145 days POS, 17/64 (26.6%) of sera had an OD value against N that was below this threshold and suggests a complementary or alternative SARS-CoV-2 antigen is needed to improve the determination of previous virus exposure in the context of vaccination for individuals infected >6 months previously.

### Neutralizing antibody responses are maintained up 10 months post onset of symptoms

The longevity of the neutralizing activity in patient sera was measured using HIV-1 based virus particles pseudotyped with SARS-CoV-2 Wuhan Spike (referred to as wild-type, WT) (**Figure 1G and Figure S1A**). Our previous study had shown a decline in neutralizing antibody titre in the first 3 months following SARS-CoV-2 infection but whether the titre would reach a steady level was not determined. The neutralization potency of matched longitudinal sera collected at timepoints up to 305 days POS revealed that the rate of decline in neutralization activity slowed in the subsequent 4–7-month period and neutralizing activity could readily be detected in 18/19 of sera tested at TP5 with a geometric mean titre (GMT) of 640. ELISA OD values for S IgG, RBD IgG and N IgG correlated well with ID_50_ of neutralization (**Figure S1B**). A cross-sectional analysis of all the wave 1 sera showed the GMT at TP3, TP4, and TP5 decreased from 1,199 to 635 and 640, respectively. The percentage of donors displaying potent neutralization (ID_50_ >2,000) was 48.2% at peak neutralization (as previously determined in Seow et al^8^) and this decreased to 27.8 %, 13.8% and 15.8 % at TP3, TP4 and TP5, respectively (**Figure S1C**).

We had previously observed that individuals experiencing the most severe disease had higher peak neutralization titres.^8^ In concordance with this, we observed higher mean peak ID_50_ values for those with most severe disease, as well as higher titres at TP3, TP4 and TP5, although this trend was not always statistically significant (**Figure 1H**). A wider heterogeneity in the magnitude of the neutralizing antibody response in the 0-3 severity group was seen at all time points studied compared to the 4-5 severity group.

Overall, neutralizing antibody response following SARS-CoV-2 infection can persist for as long as 10 months POS.

### Sera from individuals infected during UK wave 1 shows cross-neutralizing activity against SARS-CoV-2 VOCs

Initially, longitudinal sera collected from 14 individuals between days 6 and 305 POS were used to compare the magnitude and kinetics of neutralizing activity against the SARS-CoV-2 variants; B.1.1.7, P.1 and B.1.351 (**Figure 2A**). The kinetics of neutralizing activity in sera were similar against all four variants and a peak in neutralization was observed around 3-5 weeks POS followed by decline to a steady level of neutralization (**Figure 2B**).

**Figure 2:**
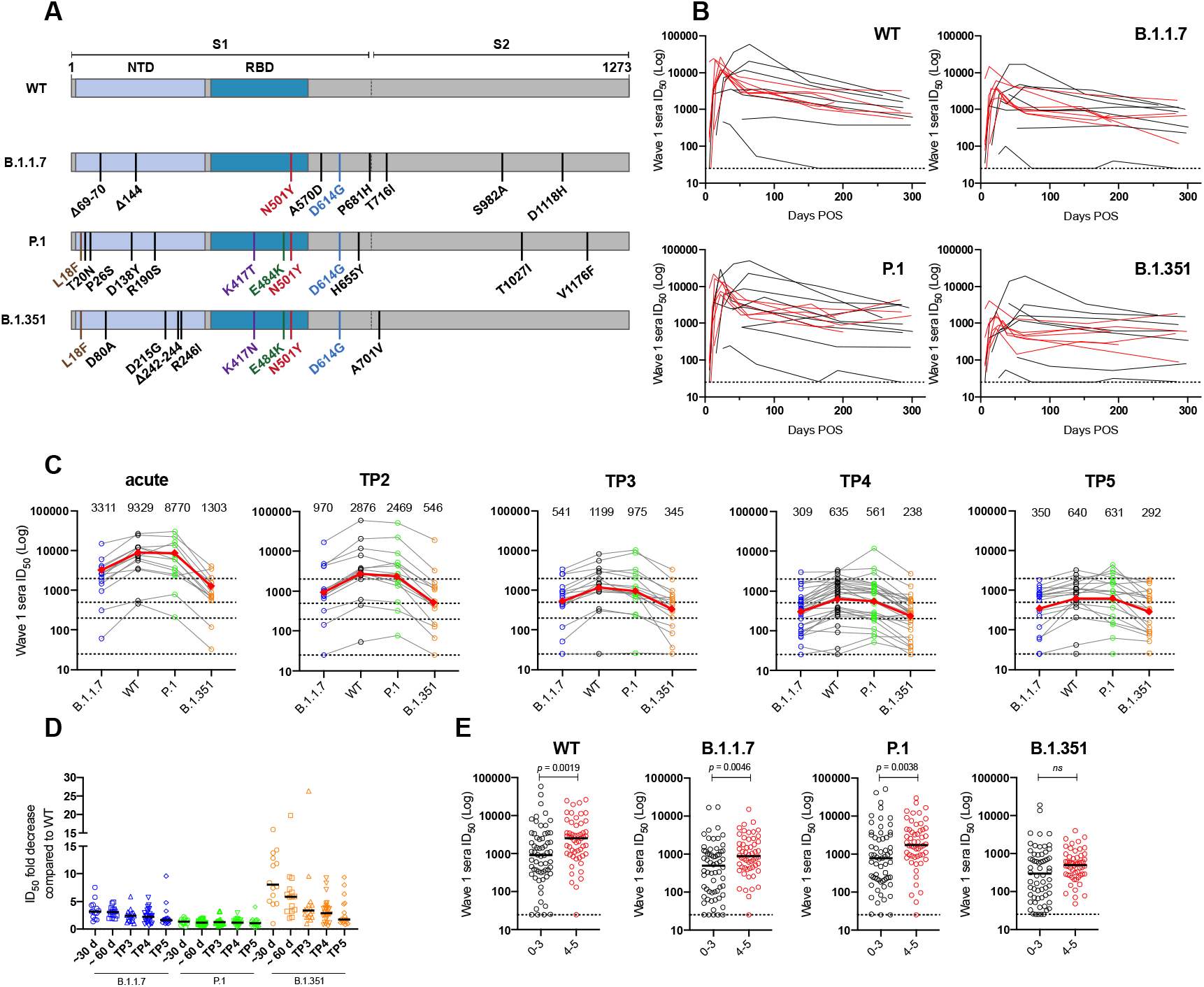
Sera from Wave 1 shows cross-neutralization of SARS-CoV-2 variants of concern. A) Schematic showing the position of Spike mutations in B.1.1.7, P.1 and B.1.351. The major Spike domains are indicated. B) Longitudinal neutralization by wave 1 sera against WT, B.1.1.7, P.1 and B.1.351. Neutralization is shown for 14 individuals. C) Neutralization of sera collected within five different time periods against the four SARS-CoV-2 variants. Geometric mean titres (GMT) against each virus are shown on each panel. Each line represents one individual, and each individual is sampled ≤1 at each timepoint. The dotted lines represent the neutralization cut-offs used to determine no, low, medium, high and potent neutralization (See **Figures S1C**). Red line represents the geometric mean titre against that virus. D) Fold change in neutralization compared to WT pseudovirus at the five timepoints. Black lines represent the average fold change. E) Comparison of the geometric mean titre between those with 0-3 (Black) and 4-5 (red) disease severity for the four variants. All sera collected up to 305 days POS are included in this analysis (n = 107). *p*-values were calculated using a Mann–Whitney two-sided test *U*-test. ns, not significant. The line represents the geometric mean ID_50_ for each group.

Having observed similar kinetics in the neutralization of VOCs, we focused further on the extent of cross-neutralizing activity of wave 1 sera collected at later time-points (145-305 days POS). Neutralization titres (ID_50_) against the four variants were measured (n = 66) and the fold change in ID_50_ compared to wild-type for each variant was compared within five time windows; acute (20-40 days POS), TP2 (55-100 days POS), TP3, TP4 and TP5 (**Figure 2C**). Neutralization potency against the P.1 variant was most similar to neutralization potency against wild-type virus at all five time-points, with an average reduction in ID_50_ ranging from 1.2-1.3 fold (**Figure 2D**). In contrast, and similar to previous reports,^19-27^ both B.1.1.7 and B.1.351 were more resistant to neutralization at all time points, with the greatest decrease in neutralization observed for B.1.351. At later timepoints, the mean fold change in neutralization ID_50_ for both the B.1.1.7 and B.1.351 variants compared to wild-type ID_50_ was decreased in magnitude (**Figure 2D**), suggesting continued antibody maturation and improved tolerance to Spike mutations. For example, the average fold reduction in ID_50_ against B.1.351 was 8.9-fold in the acute phase and this decreased to 2.9-fold at TP5. Individuals experiencing more severe COVID-19 (severity 4-5) consistently showed higher neutralization titres against the VOCs compared to those experiencing milder disease (severity 0-3) (**Figure 2E**).

Overall, wave 1 sera showed neutralizing activity against P.1, B.1.1.7 and B.1.351, albeit at a lower potency for B.1.1.7 and B.1.351.

### Sera from individuals infected with the B.1.1.7 variant retain neutralizing activity against early variants

During the UK second wave of COVID-19 in December 2020 – February 2021, the predominant variant infecting patients at St Thomas Hospital in London was B.1.1.7. Whole genome sequencing was used to confirm infection with this lineage and corresponding sera samples (n = 81) were collected from 39 individuals between 4- and 79-days POS at multiple time-points where possible. Homologous neutralization and cross-neutralizing activity were measured against wild-type, P.1 and B.1.351 pseudotyped particles (**Figure 3 and Figure S2**).

**Figure 3:**
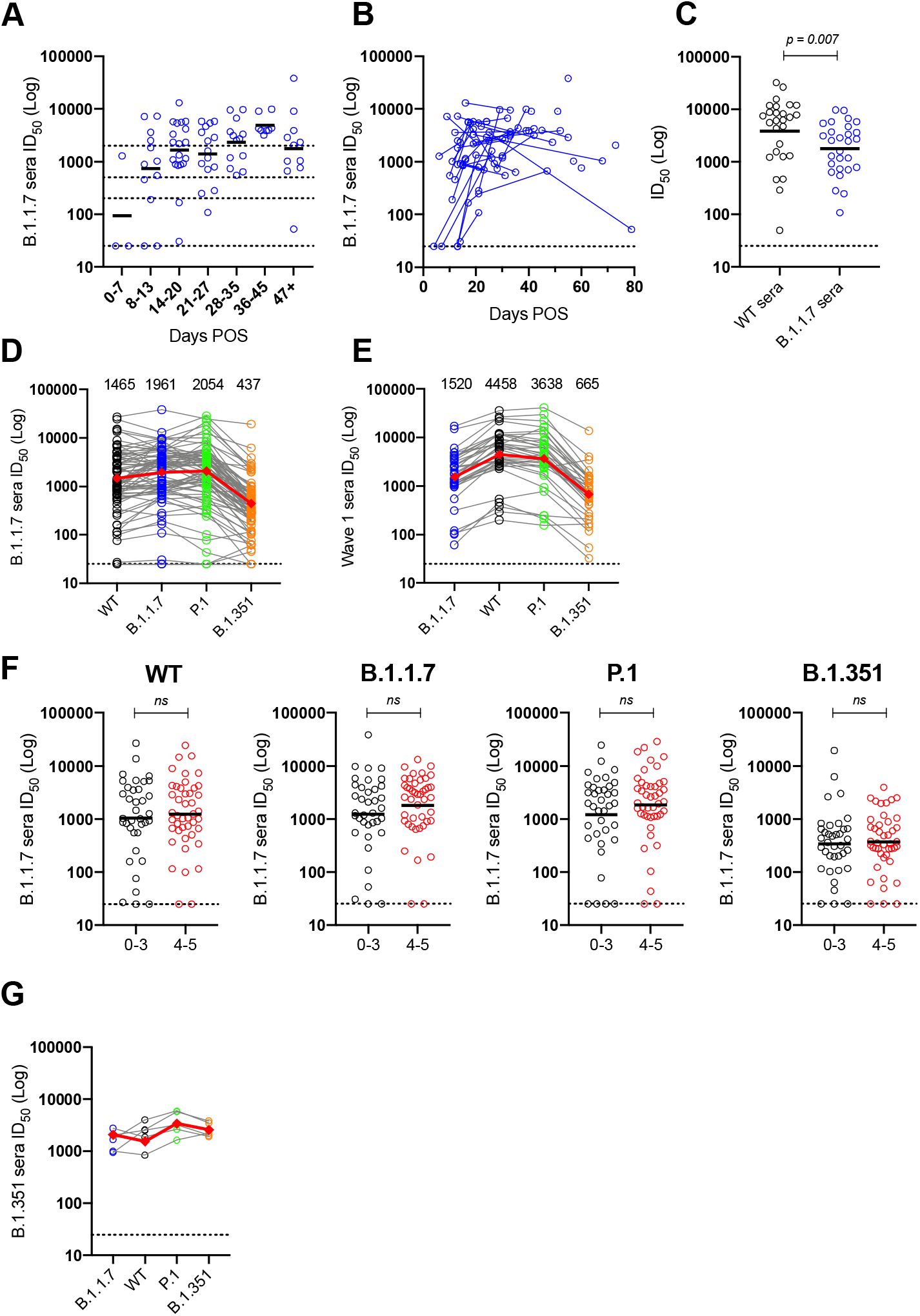
Neutralizing antibody response in individuals infected with B.1.1.7. A) Serum neutralization against B.1.1.7 at different time windows. Black line represents the geometric mean titre. B) Neutralization of B.1.1.7 pseudovirus by sequential serum samples. Each line represents samples from 1 donor across multiple timepoints. C) Comparison of homologous neutralization (i.e. neutralization of WT pseudovirus by wave 1 sera and neutralization of B.1.1.7 pseudovirus by sera from B.1.1.7 infected individuals) at peak neutralization (21-35 days POS). Line represents the geometric mean titre. *p*-values were calculated using a Mann–Whitney two-sided test *U*-test. D) Cross-neutralizing activity of sera collected between days 10-60 POS from individuals infected with B.1.1.7 against 4 SARS-CoV-2 variants (n = 74). Each line represents a serum sample. Red line represents the geometric mean titre against that virus. E) Cross-neutralizing activity of sera collected between days 10-60 POS from individuals infected in wave 1 against 4 SARS-CoV-2 variants (n = 35). Each line represents a serum sample. Red line represents the geometric mean titre against that virus. F) Comparison of the neutralization potency of B.1.1.7 sera against SARS-CoV-2 variants between individuals experiencing disease severity 0-3 and 4-5. The black lines represent the geometric mean titres. *p*-values were calculated using a Mann–Whitney two-sided test *U*-test. ns, not significant. G) Cross-neutralizing activity of sera collected from three individuals infected with B.1.351. Sera were collected at two time points from two of these individuals (between 26-52 days POS). Red line represents the geometric mean titre against that virus.

Sera from individuals infected with B.1.1.7 showed potent homologous neutralization (**Figure 3A**). Analysis of both serially collected samples (**Figure 3B**) and cross-sectional samples (**Figure 3A**) showed that the neutralization of the B.1.1.7 variant followed a similar kinetics with highest neutralization titres being detected around 3-5 weeks POS. For sera collected near the peak of the antibody response (21-35 days POS), more potent homologous neutralization was observed for wave 1 than B.1.1.7 sera (**Figure 3C**), i.e. a higher GMT ID_50_ was observed for wave 1 sera against WT pseudotyped particles compared to B.1.1.7 sera against B.1.1.7 pseudotyped particles. This may be indicative of a higher immunogenicity of the WT Spike compared to the B.1.1.7 Spike, or of increased administration of immunosuppressive drugs, e.g. Dexamethasone during the 2^nd^ wave of COVID-19 in the UK.

The majority of B.1.1.7 sera showed cross-neutralizing activity against the other VOCs (**Figure S2C**). Similar to wave 1 sera, the lowest cross-neutralization was observed against B.1.351 which exhibited an average 5.7-fold reduction in neutralizing activity compared to neutralization against B.1.1.7 across all samples studied. Neutralization of P.1 and WT were reduced by an average 1.2- and 1.7-fold compared the B.1.1.7. To enable a fair comparison of cross-neutralizing activity generated by infection with WT or B.1.1.7 virus, neutralization potency against the four viruses was compared for all sera collected between days 10 – 60 POS (**Figure 3D**). Both B.1.1.7 sera (**Figure 3D**) and wave 1 sera (**Figure 3E**) showed a reduction in neutralization of B.1.351 compared to homologous neutralization of WT and B.1.1.7 pseudotypes (average 5.9- and 8.3-fold, respectively). Neutralization of P.1 by either wave 1 or B.1.1.7 sera was largely unchanged (1.3- and 1.2-fold changes, respectively). However, in contrast to convalescent sera from wave 1 that had an average 3.3-fold reduction in B.1.1.7 neutralization, there was only an average 1.7-fold reduction in WT neutralization by B.1.1.7 sera suggesting that neutralization is retained against earlier lineage variants if infected with B.1.1.7.

As we had previously observed a correlate between disease severity and neutralization titre for wave 1 sera (**Figure 2E**), we similarly compared the geometric mean titres for those with 0-3 and 4-5 disease severity for all B.1.1.7 serum samples. In contrast to wave 1 sera, the sera from B.1.1.7 infected individuals experiencing 4-5 disease severity did not display such an enhanced neutralization potency compared to the less severe group which may also reflect the increased administration of immunosuppressive drugs during treatment (**Figure 3F**).

Overall, sera from individuals infected with the B.1.1.7 variant displayed potent cross-neutralizing activity.

### Sera from individuals infected with B.1.351 displays potent homologous neutralization

Lastly, sera were collected from three individuals receiving treatment for COVID-19 at St Thomas’ hospital who were confirmed to have been infected with the B.1.351 variant. All experienced severity 4 illness. Neutralization against the four variants was measured. Robust neutralization of B.1.351 was observed. Although only a very small sample size, in contrast to wave 1 and B.1.1.7 sera, neutralization of B.1.351 by these sera appeared more comparable to the other three variants (**Figure 3G**).

## Discussion

With the recent entry of SARS-CoV-2 into the human population, there is limited information on the longevity of the antibody response following natural infection or COVID-19 vaccination. Initial concerns were that the SARS-CoV-2 antibody response might mimic that of other human endemic coronaviruses, such as 229E, where antibody responses are short-lived and re-infections occur.^43,44^ However, the data presented here supports other recent studies^9,45-52^ showing that although neutralizing antibody titres decline from the initial peak response, robust neutralizing activity can still be detected in a large proportion of convalescent sera at up to 10 months POS. As IgM has been shown to facilitate neutralization,^8,53^ the initial decline in neutralization is likely in part due to the reduction in circulating serum IgM observed, as well as the death of short-lived antibody-secreting cells, with the sustained neutralizing activity therefore arising from long-lived plasma cells producing spike-reactive IgG.^3,51,54^ We observed a more notable decline in IgG to N compared to IgG to Spike which has also been observed by others^51^ and is particularly relevant when considering using IgG to N to determine prior SARS-CoV-2 infection in COVID-19 vaccination studies. Further assessment of the longevity of the neutralizing antibody response arising from SARS-CoV-2 natural infection will become increasingly difficult as more of the global population receive a COVID-19 vaccine.

Although sustained neutralization against the infecting SARS-CoV-2 variant is important, efficacious cross-neutralizing activity is essential for long-term protection against newly emerging variants. As RBD has been identified as a major target for the neutralizing antibody response to SARS-CoV-2, mutations K417T/N, E484K and N501Y are of particular concern for immune evasion and have been shown to lead to resistance to some RBD specific mAbs.^25,55-58^ Additionally, mutations in NTD can also lead to neutralization resistance against NTD-specific mAbs.^20,25,59^ In this present study, the largest decrease in neutralization potency for both wave 1 (overall average 4.8-fold) and B.1.1.7 sera (overall average 5.7-fold) was observed against B.1.351 which encodes RBD mutations K417N, E484K and N501Y. Despite P.1 encoding similar RBD mutations K417T, E484K and N501Y, only a very minor decrease in neutralization potency was observed. As these two VOCs also encode a different pattern of NTD and S2 mutations, these data indicate that the RBD is not the only antigenic region responsible for reduced neutralization potency and suggests that assessment of mutational profiles throughout Spike will be important when considering immune evasion by emerging viral variants.^24^

Despite the reduction in neutralization potency seen in wave 1 sera against B.1.1.7 and B.1.351, GMT of 3,331 and 1,303 (**Figures 2C and S2C**), respectively, were still observed at the neutralization peak, and neutralization (ID_50_ >25) was detected in 17/19 and 18/19 of individuals at 257-305 days against B.1.1.7 and B.1.351. These data highlight how the polyclonal nature of convalescent sera enables antiviral functionality against mutant Spikes present in emerging viral variants. Whether the neutralizing antibody titres reported here will be sufficient to protect from infection and/or severe disease is not fully understood.^3- 6,60^ Several studies have reported a lower vaccine efficacy in locations where B.1.351 is prevalent^61,62^ whereas protection against B.1.1.7 infection has been reported in Israel following vaccination with BNT162b2^63^ and following AZD1222 in the UK.^64^ Interestingly, the differential neutralization of B.1.351 and B.1.1.7 compared to WT virus decreased at later timepoints for wave 1 sera, suggesting that antibodies present at later timepoints are better able to tolerate Spike mutations. Indeed, a study by Gaebler *et al* showed that SARS-CoV-2 monoclonal antibodies isolated 6-months POS had more somatic hypermutation and displayed a greater resistance to RBD mutations.^55^ These findings suggest that COVID-19 vaccine boosting may further increase neutralization breadth and protection against newly emerging SARS-CoV-2 VOCs.

Spikes from VOCs are being investigated as second-generation vaccine candidates to tackle the challenges associated with protection against SARS-CoV-2 emerging variants^34-37^ and therefore, studying the immune response to Spike variants in natural infection can provide insight into differential Spike immunogenicity. We show that infection with B.1.1.7 elicits a robust neutralizing antibody response against B.1.1.7, P.1 and WT variants. For the majority of donors, the ID_50_s against B.1.1.7 and WT were very similar indicating that neutralizing antibodies arising from infection with B.1.1.7 are able to maintain efficacy against previously dominant SARS-CoV-2 variants. These findings contrast with Faulkner *et al* who observed a decreased level of cross-neutralization in B.1.1.7 infected individuals.^38^ However, Faulkner *et al* used sera collected at around 11 days POS and, as discussed above, cross-neutralizing activity likely develops over time. Here we show that, similar to wave 1 sera, neutralization of B.1.351 by B.1.1.7 sera was reduced compared neutralization of B.1.1.7 and suggests the shared N501Y mutation is not sufficient to overcome the B.1.351 neutralization resistance, an independent SARS-CoV-2 lineage. A study by Moyo-Gwete *et al* demonstrated that individuals infected with B.1.351 elicited potent neutralizing antibodies against B.1.351 and P.1 but reduced titres against Wuhan-D614G variant.^39^ Cele *et al* showed that B.1.351 infection generated better cross-neutralizing activity against earlier viral variants.^26^ Although a small sample size, our data broadly support these observations and further demonstrate that B.1.351 infection elicits a robust homologous neutralizing antibody response that also cross-neutralizes other VOCs.

Previous studies of wave 1 sera comparing antibody responses in individuals experiencing different disease severities has shown that higher neutralization titres are typically observed in those experiencing more severe disease.^8,21,65-67^ Here we further show that the difference in neutralization potency decreases at later timepoints. Indeed, Vanshylla *et al* observed a more rapid initial decline in neutralizing antibody titres in those who experience severe disease.^50^ A similar analysis conducted with sera from B.1.1.7 infected individuals revealed more similar neutralizing antibody responses between the two severity groups. Whether this is related to improved disease management and increased use of immunosuppressive drugs during the UK second wave infections or is intrinsic to the B.1.1.7 Spike would need to be investigated further.

In summary, using convalescent sera from individuals infected in wave 1 or individuals infected with B.1.1.7, we show that cross-neutralizing antibodies are detected up to 10 months POS in some individuals and that infection with B.1.1.7 generates a cross-neutralizing antibody response that is effective against the parental virus. These findings have implications for optimization of COVID-19 vaccines effective at eliciting a cross-neutralizing antibody response that protects against SARS-CoV-2 viral variants.

## Methods

### Patient samples

Collection of surplus serum samples was approved by South Central REC 20/SC/0310. SARS-CoV-2 cases were diagnosed by RT–PCR of respiratory samples at St Thomas’ Hospital, London. 894 serum samples from 585 individuals were saved between 04 January 2020 and 12 March 2021. Samples obtained ranged from 8 days prior up to 319 days POS. Cases were linked to corresponding genome sequencing of viral isolates from nose and throat swabs. Some sera were previously studied in Seow *et al*^8^ as stated in the manuscript.

### Plasmids

The wild-type^8^ and B.1.1.7^20,21^ Spike plasmids were described previously. B.1.1.7 mutations introduced were ΔH69/V70, ΔY144, N501Y, A570D, D614G, P681H, T716I, S982A, D1118H. Spikes encoding the variants B.1.351 and P.1 were synthesized (Genewiz, USA) and cloned into pcDNA3.1. B.1.351 mutations introduced were L18F, D80A, D215G, Delta242-244, R246I, K417N, E484K, N501Y, D614G, A701V. P.1 mutations introduced were L18F, T20N, P26S, D138Y, R190S, K417T, E484K, N501Y, D614G, H655Y, T1027I, V1176F.

### COVID-19 severity classification

The score, ranging from 0 to 5, was devised to mitigate underestimating disease severity in patients not for escalation above level one (ward-based) care. Patients diagnosed with COVID-19 were classified as follows: (0) Asymptomatic or no requirement for supplemental oxygen; (1) Requirement for supplemental oxygen (fraction of inspired oxygen (*F*iO2) < 0.4) for at least 12 h; (2) Requirement for supplemental oxygen (*F*iO2 ≥ 0.4) for at least 12 h; (3) Requirement for non-invasive ventilation/continuous positive airway not a candidate for escalation above level one (ward-based) care; (4) Requirement for intubation and mechanical ventilation or supplemental oxygen (*F*iO2 > 0.8) and peripheral oxygen saturations <90% (with no history of type 2 respiratory failure (T2RF)) or <85% (with known T2RF) for at least 12 h; (5) Requirement for ECMO.

### Viral sequencing

Whole genome sequencing of residual nose-and-throat swab from SARS-CoV-2 cases was performed using GridION (Oxford Nanopore Technology), using version 3 of the ARTIC protocol and bioinformatics pipeline.^68^ From November 2020 all samples from inpatients were assessed for sequencing. Samples were selected for sequencing if the corrected CT value was 32 or below, or the Hologic Aptima assay was above 1000 RLU, and if there was sufficient residual sample. Sequencing was performed under COG-UK ethical approval. Lineage determination was performed using updated versions of pangolin 2.0.^69^ Samples were regarded as successfully sequenced if over 50% of the genome was recovered and if lineage assignment by pangolin was given with at least 50% confidence.

### ELISA binding to N, S and RBD

ELISAs were carried out as previously described.^8,40^ All sera were heat inactivated at 56 °C for 30 min before use. High-binding ELISA plates (Corning, 3690) were coated with antigen (N protein, S glycoprotein or RBD) at 3 μg/mL (25 μl per well) in phosphate-buffered serum (PBS), either overnight at 4 °C or for 2 h at 37 °C. Wells were washed with PBS-T (PBS with 0.05% Tween-20) and then blocked with 100 μl of 5% milk in PBS-T for 1 h at room temperature. The wells were emptied, and serum diluted at 1:50 in milk was added and incubated for 2 h at room temperature. Wells were washed with PBS-T. Secondary antibody was added and incubated for 1 h at room temperature. IgM was detected using goat-anti-human-IgM-HRP (horseradish peroxidase) (1:1,000) (Sigma, catalogue no. A6907) and IgG was detected using goat-anti-human-Fc-AP (alkaline phosphatase) (1:1,000) (Jackson, catalogue no. 109-055-098). Wells were washed with PBS-T and either AP substrate (Sigma) was added and read at 405 nm (AP) or one-step 3,3′,5,5′-tetramethylbenzidine (TMB) substrate (Thermo Fisher Scientific) was added and quenched with 0.5 M H_2_S0_4_ before reading at 450 nm (HRP). Control reagents included CR3009 (2 μg/mL), CR3022 (0.2 μg/mL), negative control plasma (1:25 dilution), positive control plasma (1:50) and blank wells. ELISA measurements were performed in duplicate and the mean of the two values was used.

### SARS-CoV-2 pseudotyped virus particle preparation

Pseudotyped HIV virus incorporating the SARS-CoV-2 Spike protein (either wild-type, B.1.1.7, P.1, B.1.351) was produced in a 10 cm dish seeded the day prior with 5×10^6^ HEK293T/17 cells in 10 ml of complete Dulbecco’s Modified Eagle’s Medium (DMEM-C, 10% FBS and 1% Pen/Strep) containing 10% (vol/vol) foetal bovine serum (FBS), 100 IU/ml penicillin and 100 μg/ml streptomycin. Cells were transfected using 90 μg of PEI-Max (1 mg/mL, Polysciences) with: 15μg of HIV-luciferase plasmid, 10 μg of HIV 8.91 gag/pol plasmid and 5 μg of SARS-CoV-2 spike protein plasmid.^70,71^ The supernatant was harvested 72 hours post-transfection. Pseudotyped virus particles was filtered through a 0.45μm filter and stored at -80°C until required.

### Neutralization assay with SARS-CoV-2 pseudotyped virus

Serial dilutions of serum samples (heat inactivated at 56°C for 30mins) were prepared with DMEM media (25uL) (10% FBS and 1% Pen/Strep) and incubated with pseudotype virus (25uL) for 1-hour at 37°C in half-area 96-well plates. Next, Hela cells stably expressing the ACE2 receptor were added (10,000 cells/25µL per well) and the plates were left for 72 hours. Infection level was assessed in lysed cells with the Bright-Glo luciferase kit (Promega), using a Victor™ X3 multilabel reader (Perkin Elmer). Each serum sample was run in duplicate and was measured against the four SARS-CoV-2 variants within the same experiment using the same dilution series.

## Data Availability

Anonymised data was used in this study and is not available for distribution outside the host organisations.

## Acknowledgments

This work was funded by; King’s Together Rapid COVID-19 Call awards to MHM, KJD and SJDN, MRC Discovery Award MC/PC/15068 to SJDN, KJD and MHM, Fondation Dormeur, Vaduz for funding equipment to KJD, Huo Family Foundation Award to MHM, KJD, MSH and SJDN, MRC Genotype-to-Phenotype UK National Virology Consortium (MR/W005611/1 to MHM, KJD and SJDN), MRC Programme Grant (MR/S023747/1 to MHM), Wellcome Trust Investigator Award 106223/Z/14/Z to MHM, NIAID Awards AI150472 and AI076119 to MHM. MSH is funded by the National Institute for Health Research Clinician Scientist Award (CS-2016-16-011). The views expressed in this publication are those of the author(s) and not necessarily those of the NHS, the National Institute for Health Research or the Department of Health and Social Care. CG, SH and HW were supported by the MRC-KCL Doctoral Training Partnership in Biomedical Sciences (MR/N013700/1). SA was supported by an MRC-KCL Doctoral Training Partnership in Biomedical Sciences industrial Collaborative Award in Science & Engineering (iCASE) in partnership with Orchard Therapeutics (MR/R015643/1). NA is funded by the Wellcome Trust PhD program Cell therapies and regenerative medicine (108874/Z/15/Z). This work was supported by the Department of Health via a National Institute for Health Research comprehensive Biomedical Research Centre award to Guy’s and St. Thomas’ NHS Foundation Trust in partnership with King’s College London and King’s College Hospital NHS Foundation Trust. This study is part of the EDCTP2 programme supported by the European Union (grant number RIA2020EF-3008 COVAB). The views and opinions of authors expressed herein do not necessarily state or reflect those of EDCTP.

Thank you to Florian Krammer for provision of the RBD expression plasmid, Philip Brouwer, Marit van Gils and Rogier Sanders (University of Amsterdam) for the Spike protein construct, Leo James and Jakub Luptak for the N protein, and James Voss and Deli Huang for providing the Hela-ACE2 cells.

## Supplementary Figures

**Figure S1:**
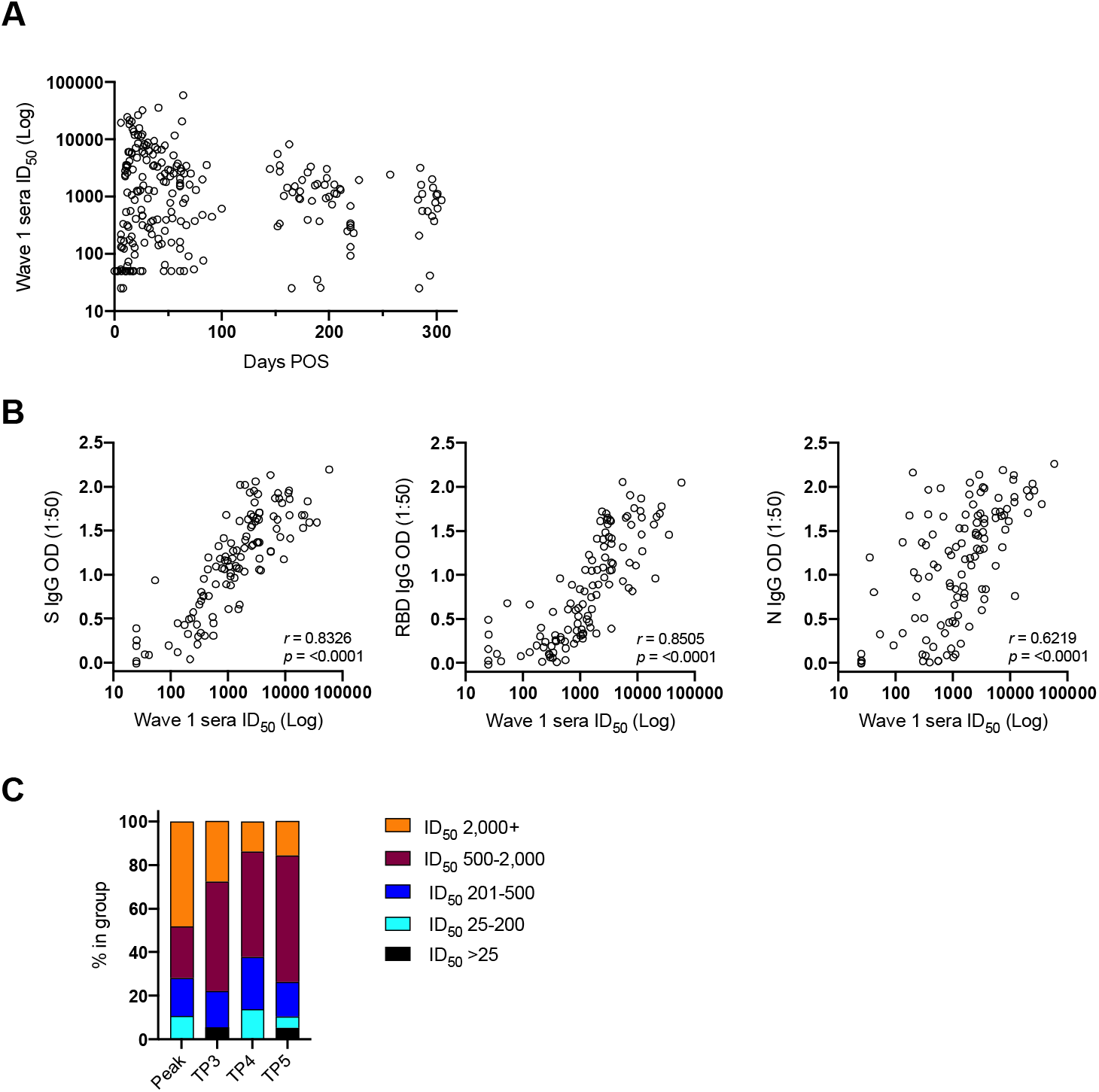
Neutralizing antibodies persist for up to 10 months post onset of symptoms. A) ID_50_ of neutralization for all wave 1 sera included in Figure 1G. B) Correlation between ID_50_ (measured against pseudovirus) and either optimal density of IgG binding to S, RBD or N. (*r*^*2*^ = 0.6942), RBD (*r*^*2*^ = 0.6250 and N protein (*r*^*2*^ = 0.3861) (Spearman’s correlation, *r*; a linear regression was used to calculate the goodness of fit, *r*^*2*^). C) Percentage of individuals in each time window with undetectable (ID_50_ <25), low (ID_50_ 25 − 200), medium (ID_50_ 201 − 500), high (ID_50_ 501 – 2,000) or potent (ID_50_ 2,000+) neutralizing antibody titres. The peak neutralization time point (n =) includes hospitalized patients and healthcare workers reported in Seow *et al*,^8^ as well as 14 additional donors reported in this study. The time point from the longitudinal samples with the peak ID_50_ was used in “peak”. TP3, TP4 and TP5 include serum samples collected between 145-175, 180-217 and 257-305 days POS.

**Figure S2:**
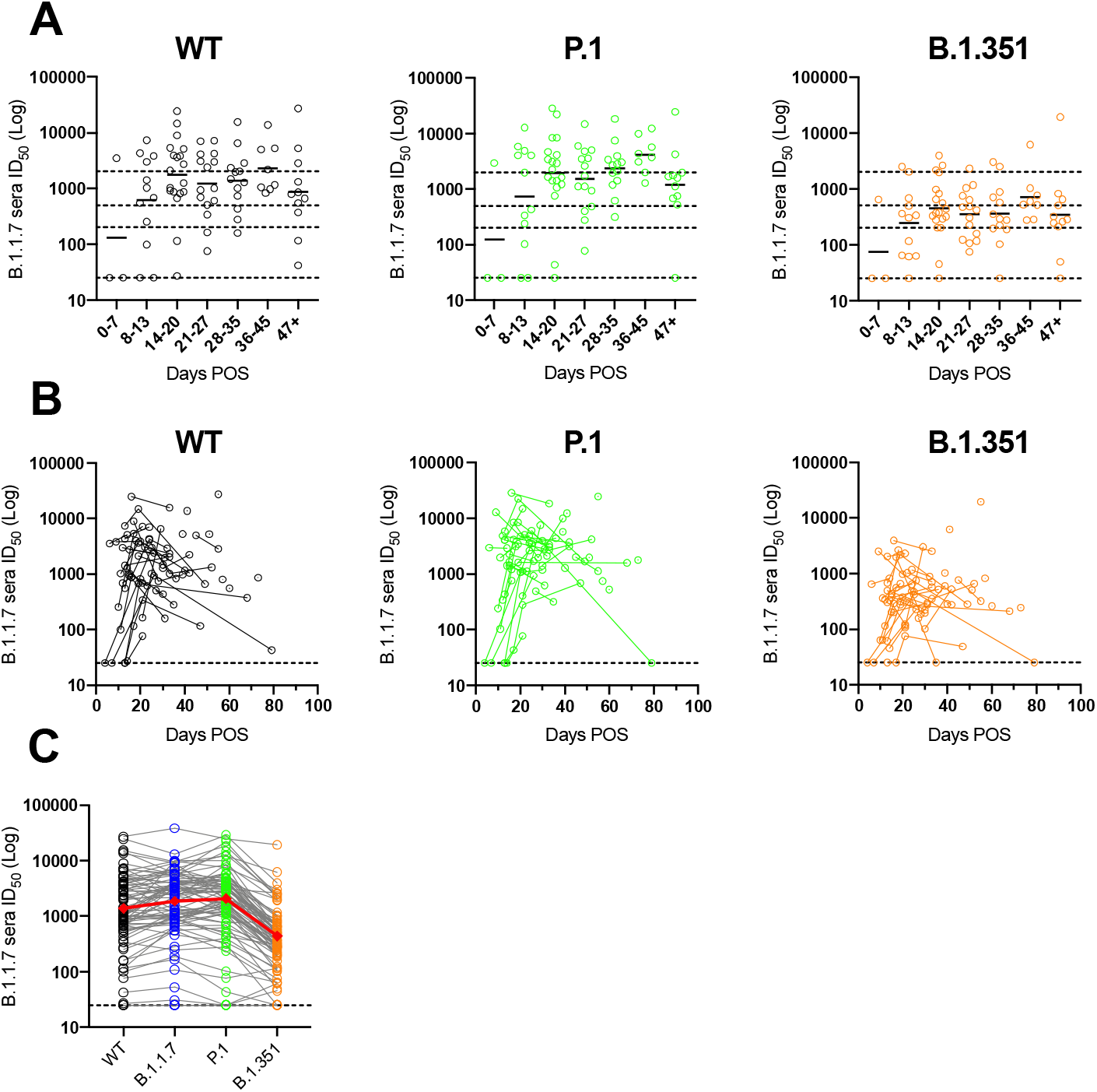
Cross-neutralizing antibody response in individuals infected with B.1.1.7. A) Serum neutralization against WT, P.1 and B.1.351 at different time windows. Black line represents the geometric mean titre. B) Neutralization of WT, P.1 and B.1.351 pseudovirus by sequential serum samples. Each line represents samples from 1 donor across multiple timepoints. C) Cross-neutralizing activity of sera from individuals infected with B.1.1.7 against 4 SARS-CoV-2 variants (n = 83). Each line represents a serum sample. Red line represents the geometric mean titre against that virus.

## Notes

### Competing Interest Statement

The authors have declared no competing interest.

### Author Declarations

Collection of surplus serum samples was approved by South Central REC 20/SC/0310.

